# Severity of Depression and Anxiety Symptoms is Reflected in Physiological and Behavioral Metrics Collected from a Consumer-Grade Wearable Ring

**DOI:** 10.64898/2026.02.06.26345566

**Authors:** Saeid Azadifar, Alireza Sameh, Laura Nauha, Mikko Kärmeniemi, Maisa Niemelä, Vahid Farrahi

## Abstract

**Background:** Modern wearable devices generate a multitude of digital metrics, which can be used to measure human function in health and disease. This study investigated whether and how digital metrics generated from a consumer-grade smart ring were different in individuals with varying severity of depression and anxiety symptoms in a large population-based cohort.

**Methods:** Cross-sectional data were obtained from the Northern Finland Birth Cohort 1986 when participants were 33–35 years of age. Participants were asked to wear the Oura Ring for two weeks, which provided digital measures of sleep architecture, nocturnal heart rate (HR), nocturnal heart rate variability indexed by the root mean square of successive differences between normal heartbeats (RMSSD), and minute-by-minute movement intensity. Participants with at least one weekday and a weekend day with no less than 15 hours of wear time were included. Two screening tools were used to assess depression and anxiety symptoms. Symptoms of anxiety were assessed using the Generalized Anxiety Disorder 7-item scale (GAD-7) and symptoms of depression and anxiety using Hopkins Symptom Checklist-25 (HSCL-25). Participants were grouped into no, mild, and moderate-to-severe anxiety symptom groups based on GAD-7, and with and without depression and anxiety symptoms based on HSCL-25. Time-series data were visualized, and statistical differences between groups were assessed using one-way ANCOVA followed by Tukey’s post hoc test.

**Results:** A total of 1,290 participants provided enough valid data and were included. Participants with moderate-to-severe anxiety assessed by GAD-7 and with depression and anxiety symptoms assessed by HSCL-25 exhibited lower percentage of rapid eye movement (REM) and deep sleep, and increased light sleep compared with the group without depression and anxiety. These two groups also showed higher HR, lower RMSSD during sleep, and lower levels of physical activity during the day. The differences between mild anxiety group with no and moderate-to-severe anxiety groups were overall less apparent on average.

**Conclusions:** Findings suggest that severity of depression and anxiety can be reflected in behavioral and physiological metrics generated by a consumer-grade wearable ring. These findings suggest that they may be a promising tool for enhancing symptoms tracking and informing personalized care strategies.

## 1. BACKGROUND

Mental health problems such as depression and anxiety impose a substantial burden on global health. These conditions are increasingly prevalent worldwide, but they are often underdiagnosed because of limited access to care, the effects of stigma, and a reliance on subjective self-reports [1, 2]. The timely diagnosis of depression and anxiety, along with proper treatment, can help manage health risks and reduce the burden on healthcare systems [3]. However, traditional diagnostic and screening approaches rely on self-report questionnaires or clinical interviews, which, although valuable, are inherently subjective and prone to recall bias [4].

Depression and anxiety are associated with changes in behavioral and cognitive factors, including alterations in sleep patterns and quality, physical activity, and psychomotor function [5]. People with mental disorders often exhibit dysregulated autonomic nervous system activity, characterized by changes in the balance between sympathetic and parasympathetic nervous system functions. These changes can potentially be associated with physiological and behavioral indicators, such as sleep, heart rate (HR), and other physical behaviors [6, 7]. Several studies have examined how physiological and behavioral changes can be used to assess symptoms of depression and anxiety [6, 8, 9]. In particular, HR and HRV have been widely studied in relation to depression and anxiety symptoms, as they may reflect changes in autonomic regulation [10–12].

The advancement of personal digital devices, especially wearable devices, has made it possible to continuously and passively capture rich streams of physiological and behavioral metrics within a person’s natural living environment. Such digital sensing signals and metrics include HR, body temperature, and physical behaviors such as active versus sedentary behavior and sleep patterns [13, 14]. The continuity of data collected via wearables is considered an advantage compared to that of laboratory-based measurements, which are typically obtained over brief periods only [15]. Recent research suggests that wearable devices can help detect changes in health status and monitor treatment responses [14, 16]. Other studies have shown that wearable-collected data, including physical activity, sleep patterns, heart rate variability, gait, and daily mobility, can be transformed into digital phenotypes and digital biomarkers that reflect health status and are associated with conditions such as cardiovascular risk, frailty, fatigue, Parkinson’s disease, and other neurodegenerative disorders [14, 17]. These approaches are particularly promising in mental research, because they can be used to non-invasively detect, assess, and predict mental health problems, including symptoms of depression and anxiety, in terms of both severity and risk [8, 14, 18].

A wide range of consumer-grade wearable devices is available today, and they are worn at various body locations depending on their design and embedded sensing technologies [14, 19]. Smart rings are a relatively new class of wearable devices that offer a lightweight alternative. They are often equipped with multiple sensors that enable the assessment and measurement of several physiological and behavioral metrics, such as HR, HRV, body temperature, physical activity, sedentary behavior, and sleep [20]. Their small size and comfortable fit make them suitable for sleep monitoring, as bulkier devices (such as smartwatches) can be uncomfortable. Research has shown that smart rings provide sleep and HR data with greater user comfort and better long-term compliance than wrist-based wearables [21]. Because they are worn on a finger, they tend to deliver more stable signals during sleep, with fewer motion artifacts [22]. These characteristics make smart rings suitable for continuous, longitudinal data collection in both research and daily life contexts, particularly with minimal interference with daily activities [21, 23].

Among all smart rings, the Oura Ring is one of the most frequently used in research and clinical studies. This multi-sensor, consumer-grade device is increasingly used for the continuous collection and screening of digital metrics and indicators to discover and validate digital biomarkers relevant to mental health issues and behavioral studies [20–22, 24]. In a pilot study using the Oura Ring, sleep HR curves and HRV changes were associated with self-reported stress and mental health indicators among young adults [25]. In another longitudinal monitoring study, the Oura Ring was used in high-burden occupational groups, such as nurses, and the results indicated strong associations between disrupted sleep cycles and worsened mental health outcomes [26]. These examples highlight the potential utility of Oura-measured digital metrics and indicators in both clinical contexts and daily life [24].

Despite growing evidence linking wearable-measured digital metrics to depression and anxiety, much remains unknown about whether and how such digital metrics vary in daily life among individuals with and without depression and anxiety in the general population [27]. This study investigates whether and how continuously and passively collected digital metrics from a consumer-grade smart ring (the Oura Ring) reflect differences in the severity of depression and anxiety symptoms in a large non-clinical population.

## 2. METHODS

### 2.1. Study population

Data for this study were obtained from the population-based Northern Finland Birth Cohort 1986 (NFBC1986) study [28]. NFBC1986 (N = 9,479) is a life-course cohort study involving participants born between July 1, 1985, and June 30, 1986, in Finland (N = 9,432 live-born) [29, 30]. Since birth, the cohort members have been regularly monitored prospectively using a broad set of clinical measurements, interviews, and postal questionnaires.

Figure 1 depicts the flow and overview of our study. We examined data from the most recent follow-up at ages 33 to 35 years. During the follow-up, cohort members residing in the city of Oulu or within a 250-km radius were invited to undergo a health examination and complete extensive health and behavior questionnaires. They were also invited to participate in sleep and physical behavior monitoring using a wearable device, specifically the Oura Ring (invited participants N = 5,717). As part of the 33–35-year follow-up, participants also self-reported their mental health symptoms using validated questionnaires to assess symptoms of depression and anxiety. Eligible participants for this study were cohort members who wore an Oura Ring and completed the mental health assessment questionnaires. The original NFBC1986 33–35-year follow-up study was approved by the Ethical Committee of the Northern Ostrobothnia Hospital District (108/2017), and written informed consent was obtained from all the participants [29].

**Figure 1:**
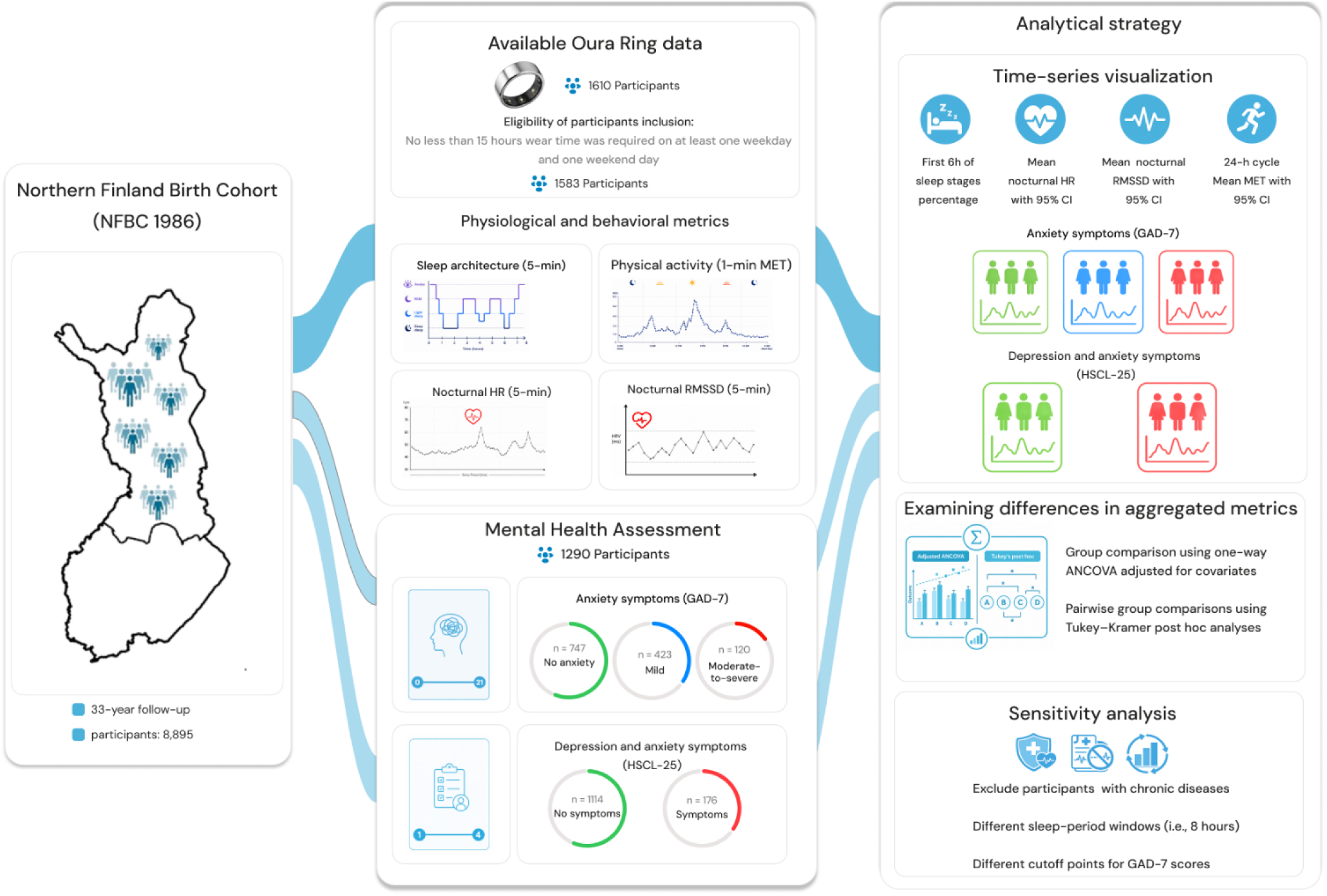
Overview of the study design, eligible participants selection, and analysis performed.

### 2.2. Measurements

#### 2.2.1 Wearable data collection with Oura Ring

Participants were asked to wear the second-generation Oura Ring (made by Ōura Health, Finland). The ring is a lightweight, waterproof, multi-sensory tracker that includes photoplethysmography (PPG) sensors for HR and respiration, a temperature sensor, and a 3D accelerometer [24, 31]. The data that the ring collects are sent automatically to a cloud server. Participants were asked to wear the ring on any finger of their non-dominant hand. Ring sizes ranged from 6 to 13 (U.S. standard), and participants were asked to wear the ring continuously for 14 days, except when using a sauna. To avoid accidental resetting of the devices, which would stop data collection, the participants were not given chargers. The ring’s battery typically lasts six consecutive 24-hour periods, according to the manufacturer. However, in the study, some ring batteries lasted up to 14 days. On average, data were collected for six full days per person. Participants did not receive any information or feedback from the ring about their sleep, activity, or behavior.

#### 2.2.2 Oura-measured digital metrics

The Oura Ring estimates the time spent awake and in light sleep, deep sleep, and rapid eye movement (REM) sleep. Sleep stages are classified using a proprietary sensor-fusion algorithm that analyzes signals derived from photoplethysmography (PPG), three-dimensional accelerometry, and peripheral skin temperature. Validation studies comparing Oura-derived sleep staging with PSG, the clinical gold standard for sleep assessment, have reported an overall good sleep staging accuracy which is comparable to those of research-grade actigraphy [23, 32]. In addition to sleep stage classification, Oura has also been validated for broader sleep parameters, including total sleep time, sleep efficiency, and sleep onset latency, although accuracy for individual non-REM sleep stages has been reported as moderate in some studies [23].

Nocturnal HR is derived from pulse wave signals captured via PPG, with inter-beat intervals (IBIs) calculated after artifact removal in Oura ring [33]. The average HR is then computed over five-minute intervals. Nocturnal HRV is quantified using the root mean square of successive differences (RMSSD) of IBIs. RMSSD is also averaged over five-minute windows. Studies have shown that HR and RMSSD values derived from the Oura Ring are strongly correlated with medical-grade electrocardiogram measurements [22, 31].

The Oura Ring estimates energy expenditure by translating accelerometer data into Metabolic Equivalent of Task (MET) values using proprietary algorithms over a 24-hour cycle. MET minutes are calculated as the product of the activity duration and its associated MET intensity [24]. A study comparing Oura-derived MET values with measurements obtained through laboratory-based calorimetry across different walking and running speeds has shown high correlation between the two methods [24].

#### 2.2.3 Mental health assessment and categorization

Depression and anxiety symptoms were assessed using two self-report screening instruments, namely the Generalized Anxiety Disorder 7-item scale (GAD-7) and Hopkins Symptom Checklist–25 (HSCL-25). The GAD-7 measures generalized anxiety symptoms severity. The HSCL-25 assesses symptoms of both depression and anxiety. These scales are widely used as reliable tools for assessing depression and anxiety in clinical and population-based research [6, 8, 34–36].

The GAD-7 consists of seven items that measure anxiety symptoms, all rated on a scale from 0 to 3, with a total possible score between 0 and 21. The instrument demonstrates high internal consistency (Cronbach’s α = 0.85) and strong criterion validity for detecting generalized anxiety disorder, with a sensitivity of 89% and specificity of 82% based on a cut-off score of 10 [37]. The GAD-7 score is commonly used to assess generalized anxiety severity, categorized as no anxiety, mild anxiety, and moderate-to-severe anxiety [37]. Following previous studies, participants were classified into anxiety severity groups based on their GAD-7 scores as follows: 0–4, no anxiety; 5–9, mild anxiety; and ≥10, moderate-to-severe anxiety [8, 37].

The HSCL-25 contains 25 items that are scored on 4-point Likert scales, where 1 means “not at all” and 4 means “extremely.” The items are divided into anxiety (10 items) and depression (15 items) symptoms [38]. In line with previous studies, a mean HSCL-25 score of ≥1.75 was considered indicative of clinically relevant depression and anxiety symptoms [39, 40].

### 2.3. Analytical strategy

We examined variations and differences in four Oura-measured behavioral and physiological metrics. The metrics were as follows: (1) five-minute sleep stages; (2) five-minute nocturnal HR; (3) five-minute nocturnal RMSSD; and (4) one-minute MET values across a 24-hour cycle (04:00 a.m. to 04:00 a.m. the following day). Although the Oura Ring provides other metrics, these four were selected because they have been used in previous studies [22, 41, 42]. Additionally, the literature suggests that these metrics could be related to mental health problems and symptoms [14, 43].

Our analysis proceeded in two phases. First, we generated time-series plots to explore the variations and differences in the trajectories of behavioral and physiological metrics among the depression and anxiety symptoms severity groups. Second, we examined differences in the aggregated duration of sleep stages, nocturnal HR, nocturnal RMSSD, and MET values across depression and anxiety symptoms severity groups using analysis of covariance (ANCOVA) with Tukey–Kramer post hoc tests to identify pairwise group differences.

#### 2.3.1 Data preparation

Sleep onset and non-wear periods were identified using the Oura Ring’s proprietary algorithm. An important step in preparing wearable data for analysis is establishing the criterion for what constitutes a valid day, as missing data due to periods of device non-wear are common in wearable studies [44]. Previous studies have used a range of minimum wear-time duration (e.g., 8, 10, 13, or 14 hours) to define a valid day, but the appropriate criterion generally depends on the type of wearable device, the nature of the data collected, and the behavioral or physiological signal being analyzed [44–46]. In the present study, a valid day was defined as a minimum of 15 hours of wear time. Because the Oura Ring is designed for continuous day-and-night wear, a higher minimum wear-time criterion than those often applied in previous studies was considered appropriate [44–46]. Adults may have different behaviors on weekdays and weekend days [47, 48]. To ensure that both weekday and weekend behavior were represented in the analyses [48], eligible participants were required to provide at least one valid weekday and one valid weekend day.

Given that each Oura-collated metric was distinct, separate data preparation procedures were applied to each. Participants had variation in both sleep onset and total sleep duration. To ensure consistency when comparing sleep architecture, nocturnal HR, and nocturnal RMSSD across participants, we standardized the analysis using a fixed sleep period. Choosing a fixed sleep period was necessary to enable comparisons of signals from sleep onset to a common endpoint across all participants. Sleep periods varied in duration from 3 to 15 hours, with most participants (n = 1,272) having at least 6 hours of recorded sleep. We used the first 6 hours of the detected sleep period to include the largest number of comparable sleep episodes. Records in which the total detected sleep duration was less than 6 hours were excluded.

Sleep stages (awake, light, deep, and REM), nocturnal HR, and nocturnal RMSSD were then extracted from the 6-hour sleep period. Sleep stages were converted to percentage to enable showing their trajectories over time. The percentage of each sleep stage was calculated for each group defined by the GAD-7 and HSCL-25 severity scales, as well as across different time points. HR and RMSSD data contained measurement gaps, including missing and zero values, likely due to poor PPG signal quality, loss of skin contact, or sensor-related errors. Since zero values for HR and RMSSD are physiologically implausible, they were treated as invalid measurements. Most participants had less than 50% missing or zero values in HR and RMSSD. Records were excluded if more than 50% of the HR or RMSSD values within the 6-hour window were missing or zero.

Movement intensities were analyzed over a 24-hour cycle. Daily one-minute MET values were arranged from 04:00 a.m. to 04:00 a.m. the following day. All non-wear periods identified by the Oura Ring’s proprietary algorithm were excluded for the analysis of 24-hour MET trajectories.

#### 2.3.2 Time-series plots of physiological and behavioral metrics

To examine variations in physiological and behavioral metrics among the depression and anxiety symptoms severity groups, we generated time-series plots of the percentages of sleep stages, nocturnal HR, nocturnal RMSSD, and MET values. These measurements were then analyzed across participant groups categorized according to GAD-7 and HSCL-25 scores. Comparisons were conducted separately for weekdays and weekend days to capture potential temporal differences in physiological regulation and behavioral rhythms. We computed the mean (with 95% confidence interval) for HR, HRV, and MET values over time in each group and displayed them in plots. For HR and RMSSD, the mean (with 95% confidence interval) was also computed for the entire analytical sample and shown in the plots to enable direct comparison with the overall sample mean.

#### 2.3.3 Examining differences in aggregated physiological and behavioral metrics

Mean sleep-stage durations, nocturnal HR, nocturnal RMSSD, and MET were averaged at the participant level separately for weekdays and weekend days. Each participant therefore contributed one observation to each ANCOVA model. Before inferential analyses, the distributions of all continuous outcome variables were assessed using histograms and quantile–quantile (Q–Q) plots. Mean durations of light, deep, and REM sleep, as well as mean nocturnal HR and MET values, were approximately normally distributed. In contrast, mean awake-stage duration and RMSSD were positively skewed and were therefore transformed using the natural logarithm. The resulting log-transformed awake-stage duration and RMSSD values were used in subsequent analyses of covariance (ANCOVAs).

One-way ANCOVAs were conducted to compare mean duration of sleep stages, nocturnal HR, nocturnal RMSSD, and MET across symptom-severity groups while adjusting for relevant covariates. When the overall ANCOVA was significant (p < 0.05), Tukey–Kramer post hoc tests were performed to examine pairwise comparisons between groups. The Tukey–Kramer procedure is appropriate for multiple comparisons involving all possible pairwise group differences when group sample sizes are unequal [49, 50]. F-statistic and overall p-value from the ANCOVA models were reported to assess between-group differences, and partial Eta squared (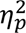) was reported as a measure of effect size.[49, 50].

Covariates were selected a priori based on their associations with both mental health symptoms and wearable-derived physiological or behavioral measures [51–54]. The covariates included sex, self-reported sociodemographic characteristics (educational attainment, employment status, marital status, and household income), lifestyle factors (smoking status and alcohol consumption), self-reported physician-diagnosed conditions (including hypertension, congenital heart disease, and type 2 diabetes), body mass index (BMI), and clinically measured cardiometabolic biomarkers (fasting glucose, triglycerides, and the low-density lipoprotein to high-density lipoprotein cholesterol ratio). To assess multicollinearity among the covariates, variance inflation factors (VIFs) were calculated. In all models, VIF values were below 5 (ranged from 1.1 to 4.7), indicating that there was no evidence of problematic multicollinearity [55]. Homogeneity of regression slopes was assessed by testing interaction terms between symptom group and each continuous covariate. No significant interactions were observed between symptoms groups and the continuous covariates (all p > 0.05) for ANCOVA model. All statistical analyses were performed using Python (version 3.11) with the *statsmodels.stats* package [56].

### 2.4. Sensitivity analysis

Three sensitivity analyses were performed. Digital metrics measured by the Oura Ring could potentially be influenced by the presence of different health conditions. To ensure that the results were not driven by such conditions, we repeated the analyses after excluding participants with any self-reported chronic diseases, including hypertension, congenital heart disease, type 1 or 2 diabetes, hypothyroidism, sleep apnea, asthma, chronic lung disease (e.g., chronic obstructive pulmonary disease (COPD)), and fibromyalgia. These conditions are known to potentially influence physiological measures such as HR and RMSSD and behavioral metrics including sleep and MET values [57, 58].

In our main analyses, we used a fixed sleep window (i.e., 6 hours) to analyze sleep stages. This approach may have resulted in the exclusion of REM sleep occurring later in the sleep period, considering that REM sleep typically occurs during the later part of the sleep period [59]. To assess the robustness of our findings with respect to REM sleep distribution, we repeated the sleep stage analyses by extending the sleep window to 8 hours.

In our main analysis, we used the most commonly cutoff values for GAD-7 score to define generalized anxiety disorders. However, there are studies that have used slightly different cutoff values. Finally, to assess the sensitivity of our findings to the cutoff values used in the main analysis, we repeated the analyses using two additional GAD-7 cutoff values reported in earlier research. Specifically, anxiety symptoms were redefined using GAD-7 scores > 4 and GAD-7 scores > 9 [37, 60, 61].

In all three sensitivity analyses, we redrew the associated plots to evaluate whether the observed patterns and associations remained consistent.

## 3. RESULTS

### 3.1. Eligible participants and descriptive statistics

In total, Oura Ring data were available for 1,610 participants. After excluding participants without valid questionnaire responses or sufficient Oura Ring data, the final analytical sample comprised 1,290 participants. Participants contributed a mean of 5.66 valid monitoring days (standard deviation (SD) 1.13; range, 2–7 days). The mean daily wear time was 23.35 hours (SD 1.20; range, 15.56–24.00 hours), indicating that participants wore the ring for approximately 23.4 hours per valid monitoring day, on average. Their mean age was 33.8 years (SD = 0.6), and 62.7% of the participants were female. Most participants held a polytechnic or university degree (58%) and were employed (82.2%). Most were married or were cohabiting (78.7%), and over half (53.6%) reported annual household incomes ranging from €50,000 to €100,000. Current smokers accounted for 18.8% of the sample, and alcohol consumption averaged 2.9 g/day (SD = 3.5). Doctor-diagnosed hypertension was reported by 1.9% of participants, congenital heart disease by 2.1%, and type 2 diabetes by 0.8%. The mean fasting glucose level was 5.0 mmol/L (SD = 0.8), and the average BMI was 26 kg/m² (SD = 4.9). Based on the GAD-7, 120 participants were categorized as having moderate-to-severe anxiety symptoms, 423 as having mild anxiety symptoms, and 747 as having no anxiety symptoms. Based on HSCL-25, 176 participants were categorized as having depression and anxiety symptoms, 1,114 participants were categorized as no depression and anxiety symptoms. The characteristics of the participants are presented in Table 1.

**Table 1:** Characteristics of all participants who had sufficient Oura Ring data and valid GAD-7 and HSCL-25 scores.

| Variables | Whole sample<br>(n = 1,290) | GAD-7 groups |  |  | HSCL-25 groups |  |
| --- | --- | --- | --- | --- | --- | --- |
|  |  | No anxiety<br>(n = 747) | Mild anxiety<br>(n = 423) | Moderate-to-severe anxiety<br>(n = 120) | No depression and anxiety<br>(n = 1,114) | Depression and anxiety<br>(n = 176) |
| **Demographics** |  |  |  |  |  |  |
| Age, yr | 33.8 (0.6) | 33.8 (0.6) | 33.8 (0.6) | 33.9 (0.6) | 33.8 (0.6) | 33.8 (0.5) |
| Sex |  |  |  |  |  |  |
| Female | 809 (62.7%) | 426 (57.0%) | 294 (69.5%) | 89 (74.2%) | 677 (60.8%) | 132 (75.0%) |
| Education |  |  |  |  |  |  |
| Comprehensive school | 102 (7.9%) | 54 (7.2%) | 37 (8.7%) | 11 (9.2%) | 75 (6.7%) | 27 (15.3%) |
| Vocational/college level | 440 (34.1%) | 247 (33.1%) | 142 (33.6%) | 51 (42.5%) | 368 (33.0%) | 72 (40.9%) |
| Polytechnic/university degree | 748 (58.0%) | 446 (59.7%) | 244 (57.7%) | 58 (48.3%) | 671 (60.3%) | 77 (43.8%) |
| Employment status |  |  |  |  |  |  |
| Employed | 1,060 (82.2%) | 642 (85.9%) | 330 (78.0%) | 88 (73.3%) | 935 (84.4%) | 121 (68.8%) |
| Unemployed | 68 (5.3%) | 26 (3.5%) | 27 (6.4%) | 15 (12.5%) | 47 (4.2%) | 21 (11.9%) |
| Other (e.g., student, homemaker) | 162 (12.5%) | 79 (10.6%) | 66 (15.6%) | 17 (14.2%) | 126 (11.4%) | 34 (19.3%) |
| Marital status |  |  |  |  |  |  |
| Married/cohabiting | 1,015 (78.7%) | 586 (78.4%) | 338 (79.9%) | 91 (75.8%) | 890 (79.9%) | 125 (71.0%) |
| Divorced/widowed | 48 (3.7%) | 23 (3.1%) | 18 (4.3%) | 7 (5.8%) | 37 (3.3%) | 11 (6.3%) |
| Unmarried | 227 (17.6%) | 135 (18.1%) | 66 (15.8%) | 22 (18.4%) | 187 (16.8%) | 40 (22.7%) |
| Household income (€ per year) |  |  |  |  |  |  |
| ≤50,000 | 509 (39.5%) | 277 (37.1%) | 175 (41.4%) | 57 (47.5%) | 414 (37.2%) | 95 (54.0%) |
| 50,001 to 100,000 | 691 (53.5%) | 409 (54.8%) | 225 (53.2%) | 57 (47.5%) | 618 (55.5%) | 73 (41.5%) |
| >100,000 | 90 (7.0%) | 61 (8.1%) | 23 (5.4%) | 6 (5.0%) | 82 (7.3%) | 8 (4.5%) |
| **Lifestyle factors** |  |  |  |  |  |  |
| Alcohol consumption, g·d<sup>-1</sup> | 2.9 (3.5) | 2.9 (3.5) | 2.6 (3.2) | 3.5 (4.4) | 2.8 (3.3) | 3.5 (4.3) |
| Smoking status |  |  |  |  |  |  |
| Nonsmoker | 417 (32.3%) | 252 (33.8%) | 127 (30%) | 38 (31.7%) | 363 (32.6%) | 54 (30.7%) |
| Former smoker | 631 (48.9%) | 352 (47.1%) | 227 (53.7%) | 52 (43.3%) | 550 (49.4%) | 81 (46.0%) |
| Current smoker | 242 (18.8%) | 143 (19.1%) | 69 (16.3%) | 30 (25.0%) | 201 (18.0%) | 41 (23.3%) |
| **Chronic disease** |  |  |  |  |  |  |
| Hypertension | 25 (1.9%) | 12 (1.6%) | 10 (2.4%) | 3 (2.5%) | 20 (1.8%) | 5 (2.8%) |
| Congenital heart disease | 27 (2.1%) | 15 (2.0%) | 10 (2.4%) | 2 (1.7%) | 22 (2%) | 5 (2.8%) |
| Type 2 diabetes | 10 (0.8%) | 2 (0.3%) | 7 (1.7%) | 1 (0.8%) | 7 (0.6%) | 3 (1.7%) |
| **Cardiometabolic biomarkers** |  |  |  |  |  |  |
| Fasting glucose, mmol·L<sup>-1</sup> | 5.0 (0.8) | 5.1 (0.9) | 5.0 (0.7) | 5.0 (0.5) | 5.0 (0.8) | 5.0 (0.8) |
| Triglycerides, mmol·L<sup>-1</sup> | 0.9 (0.6) | 1.0 (0.6) | 0.9 (0.5) | 0.9 (0.5) | 0.9 (0.6) | 0.9 (0.5) |
| Total/HDL cholesterol ratio | 3.3 (0.9) | 3.3 (0.9) | 3.2 (0.9) | 3.2 (0.8) | 3.3 (0.9) | 3.2 (0.9) |
| LDL/HDL cholesterol ratio | 2.0 (0.8) | 2.0 (0.8) | 1.9 (0.8) | 1.9 (0.8) | 2.0 (0.8) | 1.9 (0.8) |
| **Adiposity measures** |  |  |  |  |  |  |
| BMI, kg·m<sup>-2</sup> | 26.0 (4.9) | 25.9 (4.7) | 26.0 (5.2) | 26.0 (5.4) | 25.9 (4.8) | 26.5 (5.8) |
| Waist circumference, cm | 87.8 (12.9) | 88.0 (12.4) | 87.8 (13.6) | 86.9 (13.6) | 87.8 (12.7) | 88.4 (14.4) |

*Data are mean (SD) or count (%)*

### 3.2 Five-minute sleep stage patterns

Figure 2 shows the five-minute sleep stage percentage patterns across the GAD-7 and HSCL-25 symptom-severity groups. For awake percentages, the plotted mean generally appeared higher among participants with moderate-to-severe anxiety symptoms and among those with depression and anxiety symptoms, although the trajectories showed considerable overlap across groups (Figures 2a, 2e, 2i, and 2m). Light sleep stage percentage trajectories were broadly similar across the symptom-severity groups, with small visual separation over the sleep period (Figures 2c, 2g, 2k, and 2o). The deep sleep stage percentage trajectories showed a generally lower percentage among participants with moderate-to-severe anxiety symptoms and among those with depression and anxiety symptoms compared with the respective lower-symptom groups (Figures 2d, 2h, 2l, and 2p). Similarly, the plotted REM sleep stage percentage appeared somewhat lower in the higher-symptom groups during portions of the sleep period, although substantial overlap between the trajectories was observed (Figures 2b, 2f, 2j, and 2n).

**Figure 2:**
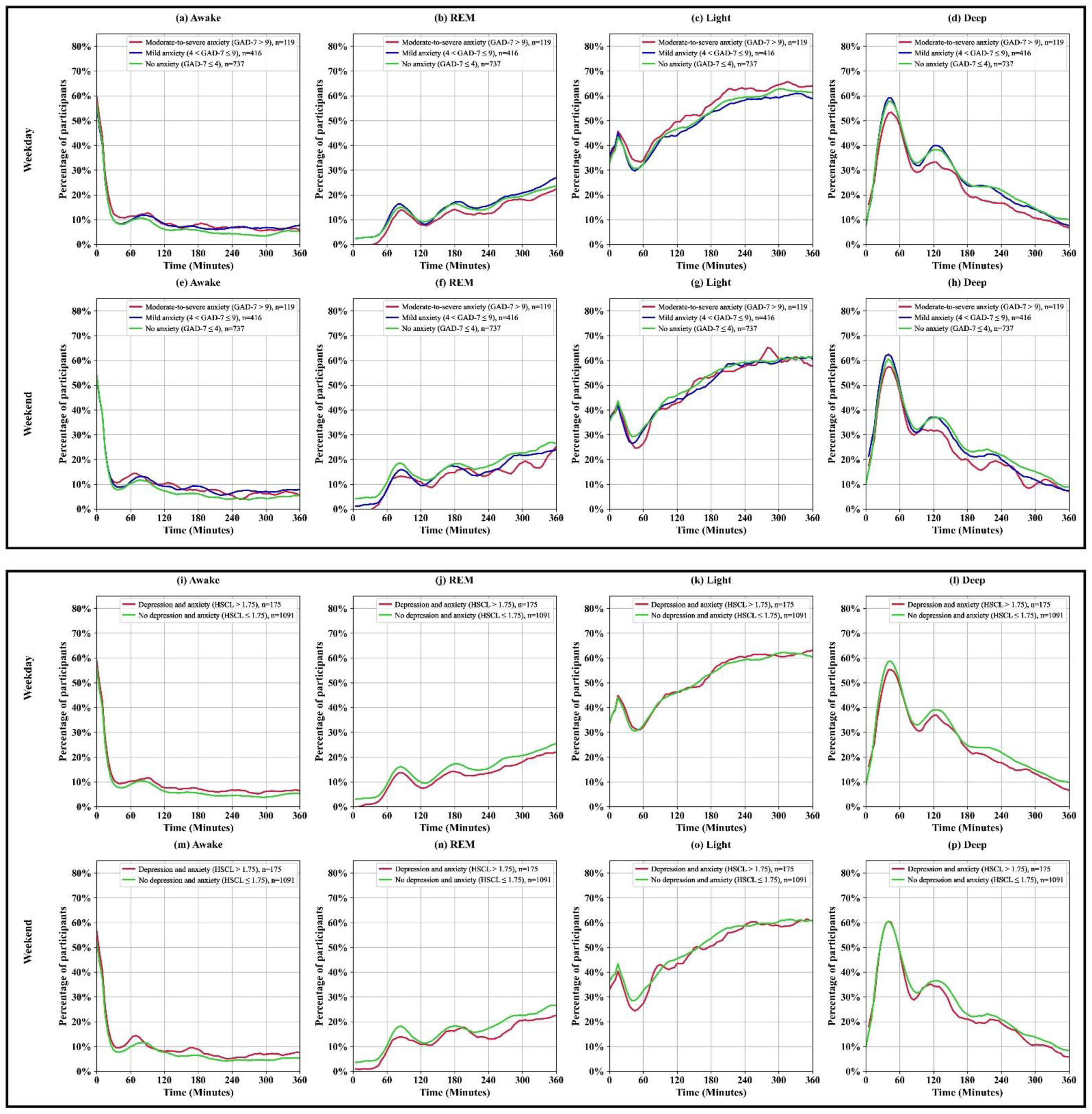
Percentage of participants in each sleep stage (Awake, Light, Deep, and REM) over time during detected sleep period on weekdays and weekend days, stratified by anxiety and depression symptoms levels. Panels (a–h) represent comparisons based on anxiety severity measured using the GAD-7 score, categorized as no anxiety, mild anxiety, and moderate-to-severe anxiety. Panels (i–p) represent comparisons based on the presence or absence of depression and anxiety symptoms measured by the HSCL-25. The top rows (a–d, i–l) show sleep stage percentages for weekdays, and the bottom rows (e–h, m–p) show percentages for weekend days. The y-axis indicates the percentage of participants, and the x-axis represents time (minutes) over the sleep period. The number of participants in each group is indicated by n in the panel legends.

### 3.3. Five-minute nocturnal heart rate patterns

Figure 3 depicts the HR patterns during the sleep period. On both weekdays and weekend days, the plotted mean of nocturnal HR appeared higher among participants with moderate-to-severe anxiety symptoms than in the whole sample, with visual separation observed across much of the sleep period (Figures 3a and 3d). The trajectories for participants with mild anxiety symptoms were more similar to the whole-sample trajectory, although the plotted mean appeared slightly higher on weekend days (Figures 3b and 3e). The no-anxiety group showed little visual separation from the whole sample on both weekdays and weekend days (Figures 3c and 3f).

**Figure 3:**
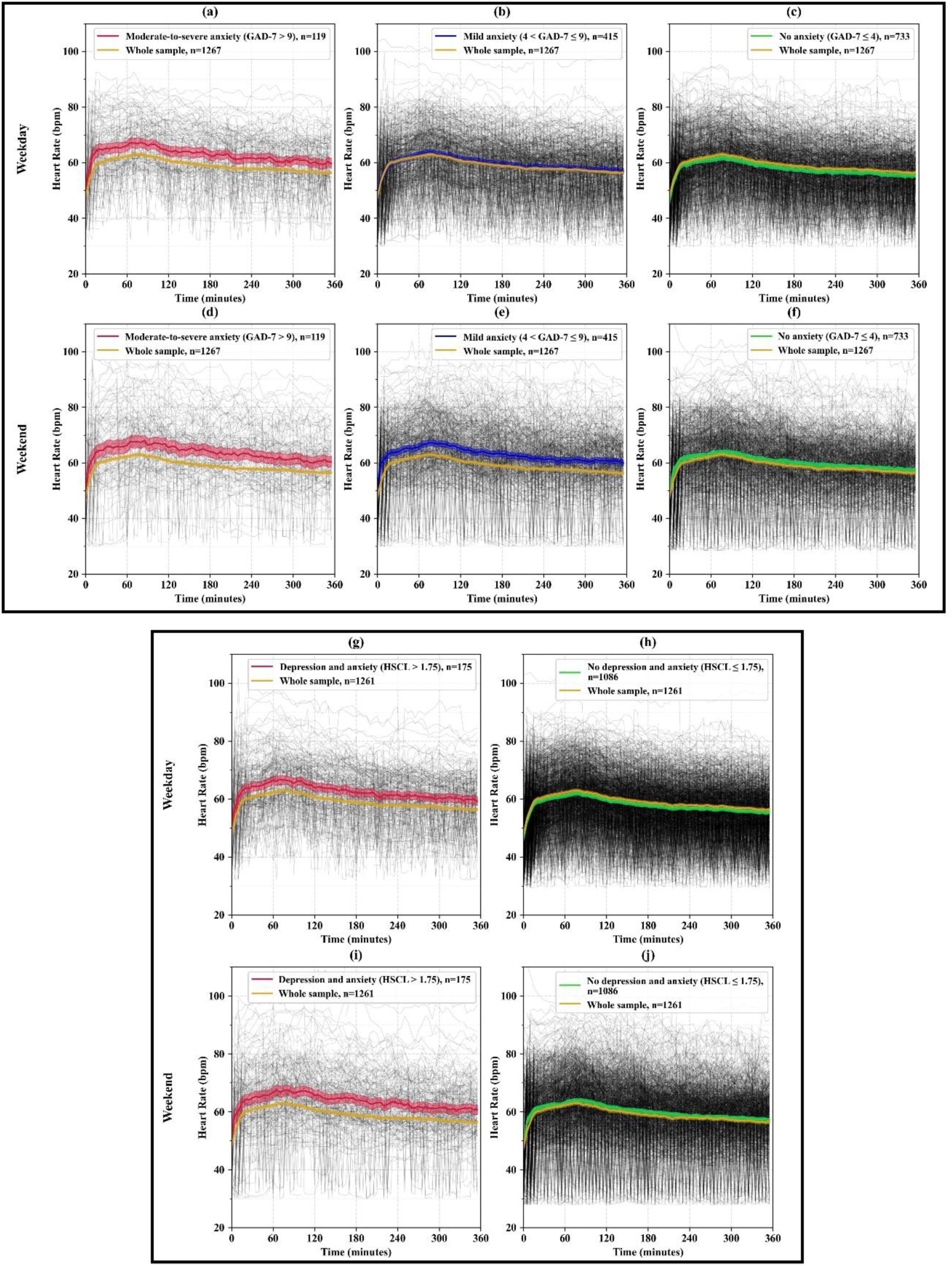
Mean HR (beats/minute) during detected sleep period on weekdays and weekend days. Panels (a–f) show the results based on anxiety severity measured by the GAD-7 scores, categorized as no anxiety, mild anxiety, and moderate-to-severe anxiety. Panels (g–j) show the results based on the no depression and anxiety and depression and anxiety symptoms measured by the HSCL-25. The number of participants in each group is indicated by *n* in the panel legends. The gray lines represent individual HR trajectories, while the colored lines indicate the mean HR for each group. The y-axis shows HR (bpm), and the x-axis represents time (minutes) over the sleep period.

When participants were categorized according to HSCL-25 scores, the plotted mean nocturnal HR appeared higher among those with depression and anxiety symptoms than in the whole sample on both weekdays and weekend days (Figures 3g and 3i). In contrast, the trajectories for participants without depression and anxiety symptoms were closely aligned with the whole-sample trajectory (Figures 3h and 3j).

### 3.4. Five-minute nocturnal RMSSD patterns

Figure 4 illustrates the RMSSD patterns during the sleep period. On both weekdays and weekend days, the plotted mean of nocturnal RMSSD appeared higher among participants with no anxiety symptoms than in the whole sample, with visual separation observed across much of the sleep period (Figures 4c and 4f). The trajectories for participants with mild anxiety symptoms were generally similar to the whole-sample trajectory (Figures 4b and 4e). Among participants with moderate-to-severe anxiety symptoms, the plotted mean nocturnal RMSSD appeared somewhat lower than the whole-sample mean, particularly on weekdays, although the trajectories showed considerable overlap (Figures 4a and 4d).

**Figure 4:**
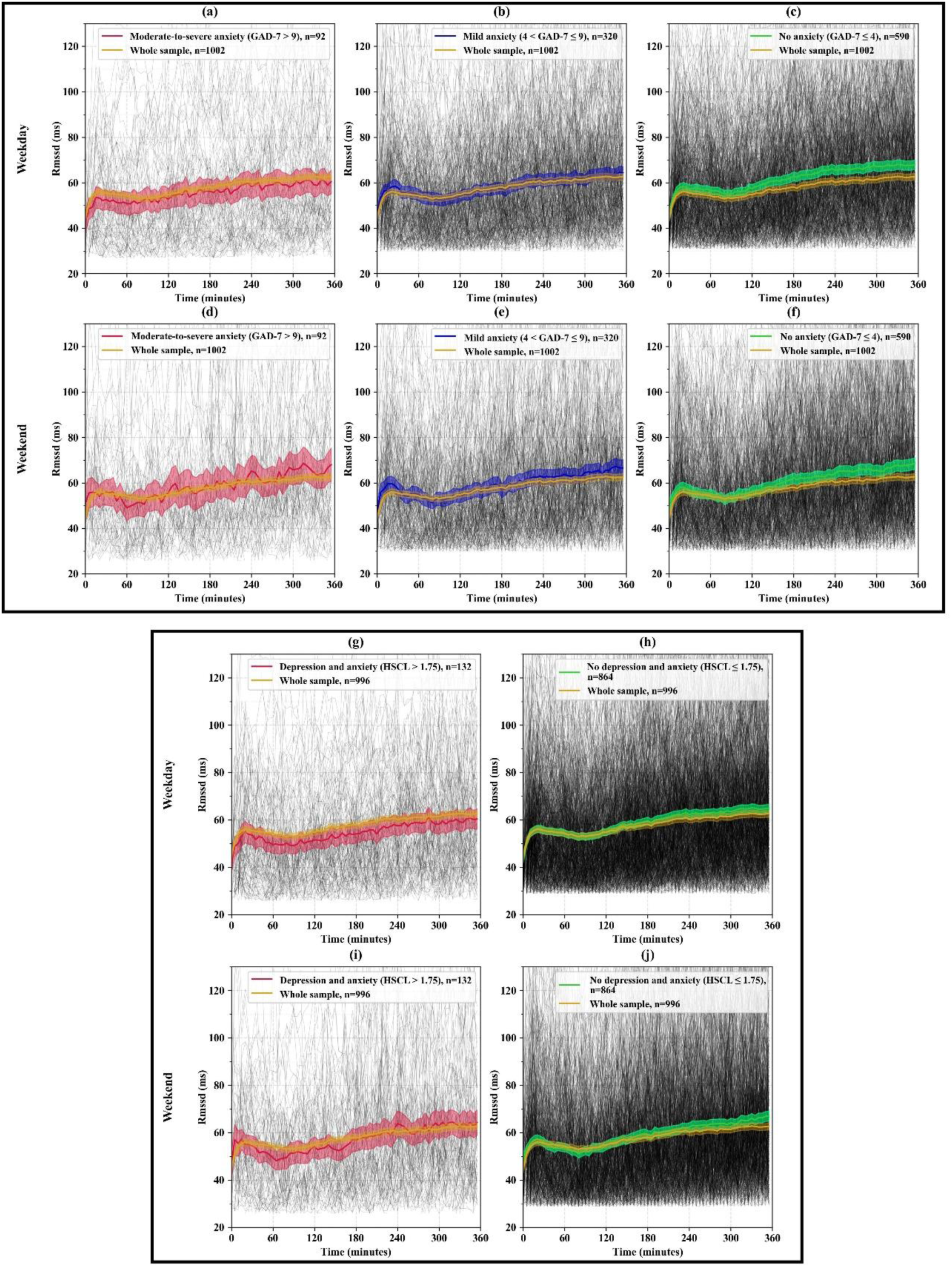
Mean RMSSD (milliseconds) during detected sleep period on weekdays and weekend days. Panels (a–f) show the results based on anxiety severity measured by the GAD-7 scores, categorized as no anxiety, mild anxiety, and moderate-to-severe anxiety. Panels (g–j) show the results based on the no depression and anxiety and depression and anxiety symptoms measured by the HSCL-25. The number of participants in each group is indicated by *n* in the panel legends. The gray lines represent individual RMSSD trajectories, while the colored lines indicate the mean RMSSD for each group. The y-axis shows RMSSD (ms), and the x-axis represents time (minutes) over the sleep period.

When participants were categorized according to HSCL-25 scores, the plotted mean nocturnal RMSSD appeared higher among those without depression and anxiety symptoms than in the whole sample on both weekdays and weekend days (Figures 4h and 4j). In contrast, participants with depression and anxiety symptoms showed a generally lower plotted mean nocturnal RMSSD than the whole sample, particularly on weekdays (Figures 4g and 4i).

### 3.5. One-minute MET activity patterns

Figure 5 shows the 24-hour profiles of one-minute average MET values. For the GAD-7 categories, participants with moderate-to-severe anxiety generally showed lower average MET levels than those with mild or no anxiety symptoms during both weekdays and weekend days (Figures 5a and 5b). The mild anxiety group generally showed values between those of the moderate-to-severe anxiety and no anxiety groups, while the no anxiety group tended to have the highest average MET levels across much of the daytime period. However, the degree of separation between groups varied over the 24-hour cycle, and the plotted confidence intervals showed some overlap.

**Figure 5:**
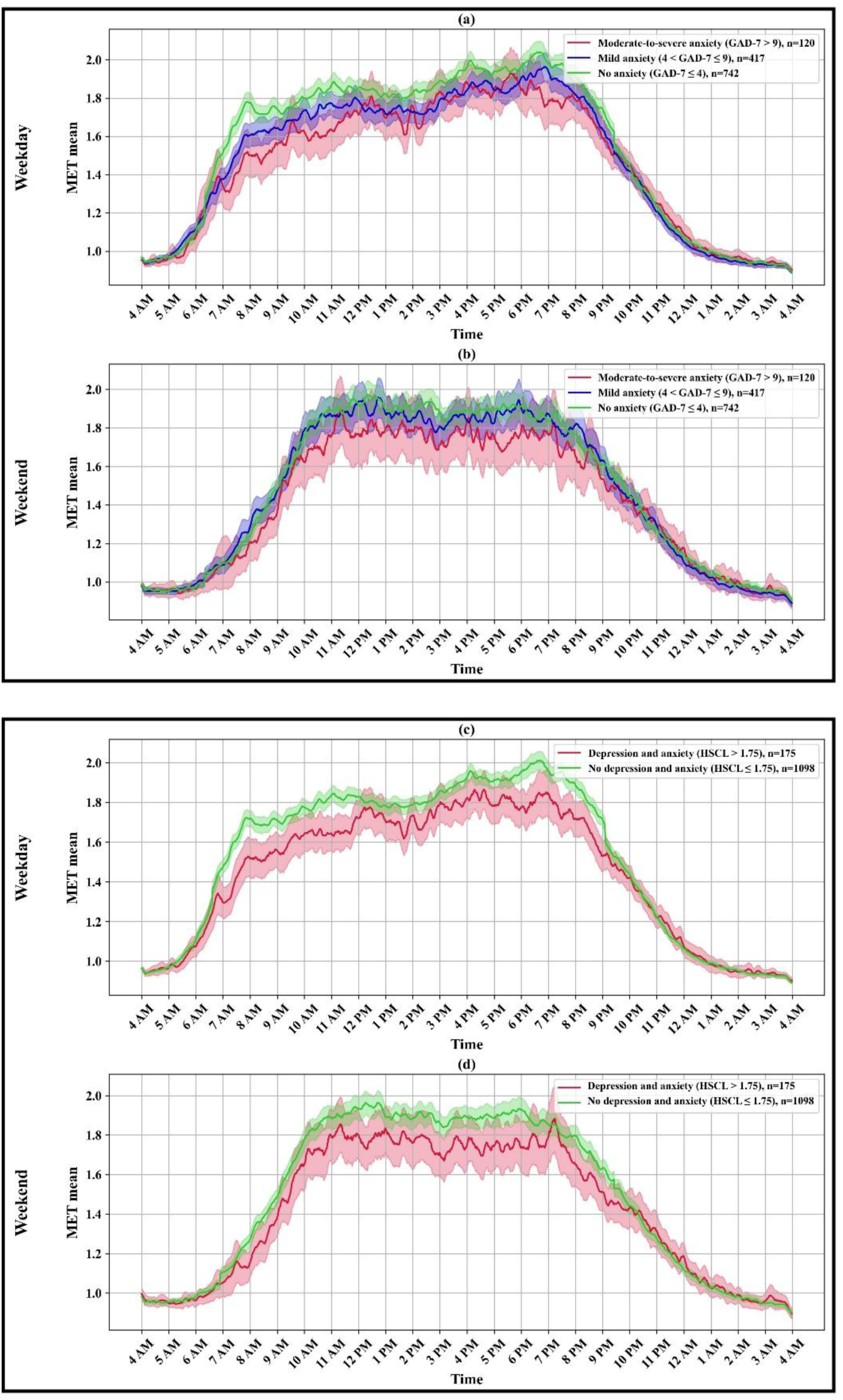
Mean MET values across 24 hours on weekdays and weekend days. Panels (a–b) show the results based on anxiety severity measured by the GAD-7 scores, categorized as no anxiety, mild anxiety, and moderate-to-severe anxiety. Panels (c–d) show the results based on the presence or absence of depression and anxiety symptoms measured by the HSCL-25. The number of participants in each group is indicated by n in the panel legends. The colored lines represent the mean MET values for each group, and the shaded areas indicate the 95% confidence intervals. The y-axis shows the mean MET values, and the x-axis represents time of day (hours).

When participants were categorized using HSCL-25 scores, those with depression and anxiety symptoms generally had lower average MET levels than those without symptoms on both weekdays and weekend days (Figures 5c and 5d). These group differences were more visible during the daytime.

### 3.6. Differences in aggregated metrics

The results of the one-way ANCOVA and Tukey–Kramer post hoc analyses are presented in Table 2. All reported means are adjusted means estimated from the ANCOVA models after controlling for the covariates. The residuals of the ANCOVA models were approximately normally distributed. For GAD-7 symptoms categories, there were statistically significant between-group differences in weekdays mean light sleep (overall p-value: 0.009), deep sleep (overall p-value: 0.020), REM sleep (overall p-value: 0.016) durations, and in mean nocturnal HR (overall p-value: 0.028), nocturnal RMSSD (overall p-value: 0.007), and MET (overall p-value: 0.025). On weekend days, statistically significant between-group differences were observed for mean light sleep (overall p-value: 0.042), deep sleep (overall p-value: 0.018), REM sleep (overall p-value: 0.028) durations, and in mean MET (overall p-value: 0.030). However, the corresponding effect sizes were small (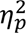 range: 0.0065-0.0095).

**Table 2:**
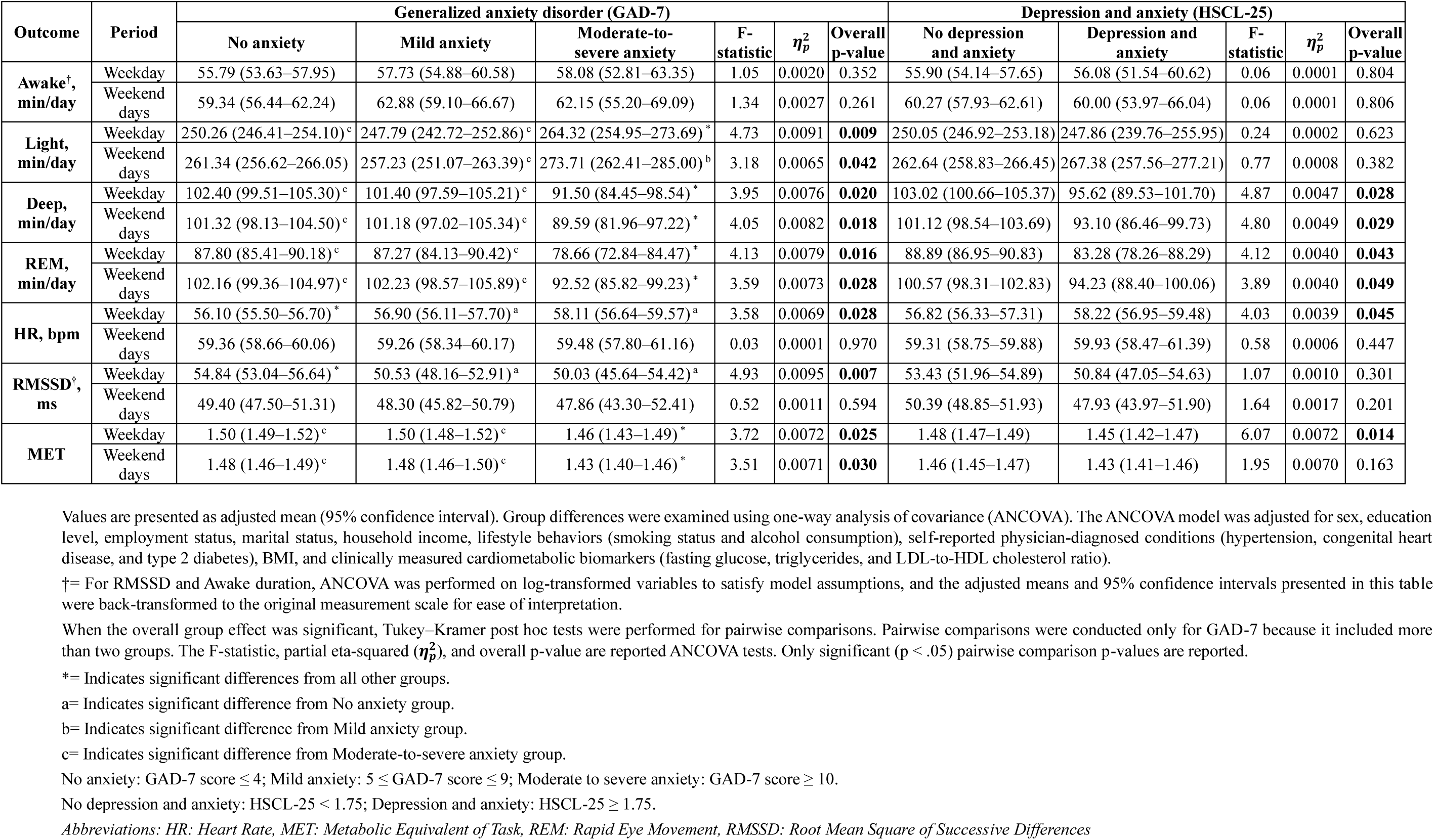
The results of one-way ANCOVA and Tukey-Kramer post-hoc tests that examined aggregated sleep stage duration (Awake, Light, Deep, *and* REM), nocturnal HR, nocturnal RMSSD, and MET values across different anxiety and depression symptoms groups.

Pairwise comparisons showed that the moderate-to-severe anxiety group had significantly higher weekday mean light sleep duration compared to the other anxiety groups (adjusted mean: 264.32 vs. 247.79 and 250.26 min/day; p < 0.05). The moderate-to-severe anxiety group also exhibited significantly lower mean deep sleep and REM sleep durations on both weekdays and weekend days compared to the other anxiety groups (91.50 vs. 101.40 and 102.40 min/day; p < 0.05), (89.59 vs. 101.18 and 101.32 min/day; p < 0.05), (78.66 vs. 87.27 and 87.80 min/day; p < 0.05), and (92.52 vs. 102.23 and 102.16 min/day; p < 0.05), respectively. In addition, the moderate-to-severe anxiety group showed higher mean HR during weekdays and lower mean MET during both weekdays and weekends compared to the other anxiety groups. Conversely, participants with no anxiety symptoms demonstrated lower weekday mean HR, higher weekday mean RMSSD, and higher weekday mean MET compared to the other anxiety groups, respectively.

For HSCL-25 symptoms categories, participants with depression and anxiety symptoms showed significantly lower mean deep sleep duration on both weekdays compared with those without these symptoms (95.62 vs. 103.02 min/day; overall p-value: 0.028) and on weekend days (93.10 vs. 101.12 min/day; overall p-value: 0.029). Mean REM sleep was also significantly lower in the depression and anxiety symptoms group on weekdays (83.28 vs. 88.89 min/day; overall p-value: 0.043) and weekend days (94.23 vs. 100.57 min/day; overall p-value: 0.049). Weekday nocturnal HR differed significantly between HSCL-25 groups, with participants with depression and anxiety symptoms showing higher weekday mean nocturnal HR (58.22 vs. 56.82 bpm; overall p-value: 0.045). Weekday mean MET was lower among participants with depression and anxiety symptoms (1.45 vs. 1.48; overall p-value: 0.014). However, the corresponding effect sizes were small (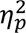 range: 0.0039-0.0072).

The results of all sensitivity analyses are presented in Additional file 1. After participants with chronic diseases were excluded, 962 participants remained in the analysis. The characteristics of the remaining participants are presented in Table S1. The overall patterns of association observed in the primary analyses remained broadly similar after this exclusion (Figures S1–S4). Similarly, repeating the analyses using a fixed 8-hour sleep window yielded results consistent with the primary findings (Figure S5). A total of 1,044 participants had at least one sleep period with a minimum of 8 hours were therefore included in this analysis. The characteristics of the remaining participants are presented in Table S2. Visual patterns during the final 2 hours of sleep were similar to those observed during the first 6 hours, with between-group differences across all sleep stages remaining also evident during the final 2 hours of the sleep period.

Similarly, the use of alternative GAD-7 cutoff scores produced findings consistent with the main analyses. No noticeable differences were observed in overall patterns of findings when alternative GAD-7 cutoff scores for anxiety symptoms severity were used (Figures S6–S13).

## 4. DISCUSSION

This exploratory study provides novel evidence that digital metrics captured by a smart ring may vary with the severity of depression and anxiety symptoms. The analysis of diurnal and nocturnal time-series data, including sleep stages, nocturnal HR, nocturnal HRV (indexed by RMSSD), and MET values, showed differential patterns across groups with varying severity of anxiety and depression symptoms. Our findings show that, individuals with moderate-to-severe anxiety symptoms (assessed based on GAD-7) and with depression and anxiety symptoms (assessed based on HSCL-25) had less daytime physical activity, altered sleep architecture (e.g., reduced REM and deep sleep), higher nocturnal HR, and lower nocturnal HRV compared to the whole sample population. These differences were generally observed for both weekdays and weekend days. The patterns remained the same even after participants with chronic diseases were excluded.

Wearable-estimated sleep architecture showed differential patterns across groups with varying levels of depression and anxiety. Most apparent differences were observed for light, deep, and REM sleep stages. The duration of deep and REM sleep stages was significantly lower in the moderate-to-severe anxiety group across both weekdays and weekend days. In recent years, the trend in mental health research has shifted from static self-report assessments toward continuous data-driven monitoring aided by wearable sensing and mobile technologies [61]. Several studies have previously shown that digital metrics collected via wearable devices could potentially be used as digital biomarkers or digital phenotypes of mental health risk [62]. These works have explored whether and how objectively or continuously measured sleep parameters (e.g. sleep timing, duration, quality, and circadian rhythmicity) are related to the risk of depression and anxiety [63]. This study adds to previous research by showing that consumer-grade wearable devices may be able to detect changes in sleep architectures that are linked to the severity of depression and anxiety symptoms.

Among the many physiological biomarkers that can be collected in both laboratory and daily life, HR and HRV stand out as particularly promising noninvasive metrics for predicting and detecting psychological states [64]. They are relatively easy to measure, especially with the growing ubiquity of wearable technologies [65]. Existing studies have previously examined the possible relationships between 24-hour and resting HR and HRV with symptoms of depression or anxiety [62, 66, 67]. Some have identified distinct 24-hour and resting HR and HRV profiles that are typically observed among people with mental disorders, whereas people without such conditions do not display the same patterns [62, 68]. However, HR and HRV adjust constantly in response to physical and emotional demands on the body. During the day, HR and HRV can be physiologically high because of high levels of activity. During sleep, when there is minimal movement and physical demand, it is likely that HR and HRV can be measured by wearables with less artifact and noise, potentially serving as better physiological indicators. This point is especially relevant for depression and anxiety, which are often influenced by stress and emotional stimuli [69, 70].

The Oura Ring obtains HR and HRV data only during nocturnal sleep. Our study is one of the first to demonstrate that nocturnal HR and nocturnal HRV collected via commercial wearable devices in free-living conditions could differ across individuals with varying levels of depression and anxiety symptoms. The findings showed that, on average, participants with symptoms of depression and anxiety had higher nocturnal HR and lower nocturnal RMSSD. The RMSSD is widely used in mobile health tools as an indicator of stress and autonomic nervous system balance [71]. In addition, low HRV is an established marker of reduced adaptability to stress and often occurs in individuals with anxiety or depression [72]. One study found that individuals with severe depressive and anxiety symptoms had relatively low HRV, as measured by smartwatch sensors, over a 4-week period [73]. Other studies on wearable devices similarly found that anxiety disorders were associated with significantly reduced HRV outside of clinical settings [68]. Our findings add to the evidence that HR and HRV data, even when collected by consumer-wearable devices, can potentially serve as objective and non-invasive indicators of depression and anxiety [68, 74].

Individuals with higher depression and anxiety symptoms appeared to be, on average, less physically active during the day than people without symptoms. This pattern was more apparent during the daytime, and these activity patterns occurred on both weekdays and weekend days. The finding shows a possible link between overall lower movement intensity and higher depression and anxiety symptoms. Wearable motion and activity tracking has previously been used to differentiate people who experience depression or anxiety symptoms from those without such symptoms, especially in large population-based samples [48]. It has been previously shown that people with higher levels of depression and anxiety generally exhibit less physical activity and distinct movement patterns compared with people who lack such symptoms [6]. This data can be leveraged to develop diagnostic models for identifying individuals at risk [8]. For example, studies in which wearable devices were used to track daily activity behaviors showed that machine learning can predict depression and anxiety based on physical activity, sedentary, and sleep metrics [8, 48]. Our study showed lower average movement intensity among individuals with depressive and anxiety symptoms, compared to people without these symptoms during parts of the daytime period; however, time-specific differences were not statistically tested.

Analysis of aggregated physiological and behavioral metrics revealed statistically significant differences across symptom severity groups in sleep stage duration, nocturnal HR, nocturnal RMSSD, and MET values. Nevertheless, the observed effect sizes were small. While aggregated wearable-derived measures are commonly used in digital biomarker research and digital phenotyping, summarizing continuously collected data into average values may mask meaningful temporal variations and limit the detection of relevant patterns [75, 76]. These findings suggest that future studies should consider analytical approaches capable of leveraging the richness and temporal dynamics of physiological and behavioral signals to capture important and relevant health-related information.

### 4.1. Strengths and limitations

The consistency of the results after these exclusions shows that the patterns in sleep, cardiovascular physiology, and physical activity are probably related to mental health symptoms rather than comorbid physical illnesses. This study has several key strengths. First, we analyzed a relatively large population-based sample of adults [77]. The sample was larger than those in previous studies on using smart rings or wearable-collected data to predict and screen for depression and anxiety [78, 79]. Second, we examined a comprehensive set of behavioral and physiological metrics captured continuously in daily life, not just in a laboratory. These included sleep architecture, nocturnal HR, nocturnal RMSSD, and physical activity intensity measured via METs. A third strength lies in the psychological assessments, which were conducted using two validated instruments. These tools are widely recognized for their reliability, cross-cultural applicability, and clinical relevance in measuring symptoms of anxiety and depression [37, 80]. A fourth strength is the robustness of our findings, which was shown through sensitivity analyses. These analyses excluded participants with chronic health conditions and tested different sleep-period window sizes and different cutoff scores for anxiety symptoms. The consistency of the results after these exclusions shows that the patterns in sleep, cardiovascular physiology, and physical activity are probably related to mental health symptoms rather than comorbid physical illnesses.

Despite these strengths, limitations must be acknowledged. One of the main limitations of this study is its observational and cross-sectional design, which limits causal inference. Since the data collection and outcomes (depression and anxiety symptoms) were measured at the same time, we cannot determine the direction of the association. Therefore, longitudinal studies are needed to better understand the possible causal nature of these associations. Another limitation involves the accuracy of sleep staging, particularly when distinguishing light sleep from REM sleep using consumer-grade wearables. Although recent studies support the acceptable validity of consumer-grade wearables for sleep staging [23, 79], there has been some concerns regarding the accuracy of wearable devices in sleep staging [23]. The resolution of wearables is also limited compared to PSG [41], which is the clinical gold standard for sleep staging. Nevertheless, the ecological validity and capacity for continuous longitudinal monitoring make wearables highly practical for large-scale studies [81, 82]. In brief, although validation studies support the general accuracy of Oura sleep metrics [79, 81–83], consumer-grade devices remain an indirect proxy compared to clinical tools [82]. The proprietary nature of Oura’s algorithms could also be considered as a limitation, which restricts transparency regarding how the specific metrics (e.g., METs and sleep stage categorization) are derived. This could affect reproducibility and hinder cross-device comparisons, especially if digital metrics are compared across brands or platforms. Although other HRV indices may capture complementary aspects of autonomic regulation relevant to depression and anxiety [84], RMSSD was used in this study because it was the only HRV metric available from the Oura Ring. The time-series plots were examined descriptively, without statistical testing of differences at specific time points. We observed noticeable differences in trajectories of the wearable-collected behavioral and physiological signals across depression and anxiety symptoms groups. However, when the differences were examined using aggregated daily mean of sleep stage durations, HR, RMSSD, and MET the observed effect sizes were small. Currently, the extent to which these observed differences could translate into clinically meaningful differences at the individual level remains uncertain. Our findings should therefore be interpreted primarily as exploratory analysis rather than as evidence that smart-ring metrics can distinguish clinically relevant mental health differences between individuals. Finally, because the study included Finnish adults aged 33–35 years, the findings may not be directly generalizable to other age groups or populations. Future studies should examine these associations in more diverse populations.

## 5. CONCLUSIONS

This study provides novel evidence that depression and anxiety severity could be reflected in passive physiological and behavioral metrics captured by a consumer-grade wearable device. Groups of participants with different levels of depression and anxiety symptoms exhibited differences in sleep architecture, HR, HRV, and movement intensities. Our results reveal that groups with higher levels of depression and anxiety symptoms exhibited reduced deep and REM sleep, higher nocturnal HR, lower RMSSD, and decreased daytime MET values. Overall, our findings support the use of consumer-grade wearable devices as tools for screening and detecting depression and anxiety in daily life settings, potentially complementing existing clinical tools and methods for mental health disorders. These findings provide further support for the growing body of research on integrating wearable digital devices into broader mental health assessments, where passive data streams and digital metrics can enhance symptoms tracking and inform personalized care strategies.

## Abbreviations

ANCOVA: Analysis of Covariance
BMI: Body Mass Index
CI: Confidence Interval
COPD: Chronic Obstructive Pulmonary Disease
GAD-7: Generalized Anxiety Disorder 7-item scale
HR: Heart Rate
HRV: Heart Rate Variability
HSCL-25: Hopkins Symptom Checklist-25
IBI: Inter-beat Interval
MET: Metabolic Equivalent of Task
NFBC1986: Northern Finland Birth Cohort 1986
PPG: Photoplethysmography
PSG: Polysomnography
Q–Q: Quantile–Quantile
REM: Rapid Eye Movement
RMSSD: Root Mean Square of Successive Differences
SD: Standard Deviation
VIF: Variance Inflation Factor

## Declarations

## Acknowledgements

We wish to thank all cohort members, researchers and NFBC project center personnel who participated in the NFBC data collection.

## Author contributions

**SA** and **AS**: Conceptualization and study design, Data analysis and interpretation, Methodology development or software/tool creation, Drafting and revising the manuscript. SA and AS contributed equally to this work (first two co-authors with same contribution).

**LN**: Conceptualization, study design and revising the manuscript

**MK**: Conceptualization and revising the manuscript

**MN**: Data acquisition or provision, Conceptualization and revising the manuscript

**VF**: Conceptualization and study design, Revising the manuscript, Supervision

All authors read and approved the final manuscript.

## Ethics Approval and Consent to Participate

The original NFBC1986 33–35-year follow-up study was approved by the Ethical Committee of the Northern Ostrobothnia Hospital District (108/2017), and written informed consent was obtained from all the participants.

## Consent for Publication

Not applicable.

## Data availability

NFBC data is available from the University of Oulu, Infrastructure for Population Studies. Permission to use the data can be applied for research purposes via electronic material request portal. In the use of data, we follow the EU 395 general data protection regulation (679/2016) and Finnish Data Protection Act. The use of personal data is based on cohort participant’s written informed consent at his/her latest follow up study, which may cause limitations to its use. Please, contact NFBC project center NFBCprojectcenter(at)oulu.fi) and visit the cohort website (www.oulu.fi/nfbc) for more information. All analyses presented in this manuscript were performed in a secure computing environment. The code used for the analyses can be made available upon request from the co-first author (SA) for those who have access to the data.

## Supplementary Information

**Additional file 1:** Table S1. Characteristics of participants after exclusion of those with chronic diseases. Figures S1–S4. Results of the sensitivity analysis after exclusion of participants with chronic diseases. Table S2. Characteristics of participants included in the sensitivity analysis using an 8-hour sleep-period window. Figure S5. Results of the sensitivity analysis using an 8-hour sleep-period window. Figures S6–S13. Results of the sensitivity analyses using alternative GAD-7 cutoff scores for anxiety symptom severity.

## REFERENCES

1. Ali A, King M, Strydom A, Hassiotis A. Self-reported stigma and symptoms of anxiety and depression in people with intellectual disabilities: Findings from a cross sectional study in England. J Affect Disord. 2015;187:224–31.

2. Cook S, Kudryavtsev A V., Bobrova N, Saburova L, Denisova D, Malyutina S, et al. Prevalence of symptoms, ever having received a diagnosis and treatment of depression and anxiety, and associations with health service use amongst the general population in two Russian cities. BMC Psychiatry. 2020;20:1–11.

3. Chisholm D, Sweeny K, Sheehan P, Rasmussen B, Smit F, Cuijpers P, et al. Scaling-up treatment of depression and anxiety: a global return on investment analysis. The Lancet Psychiatry. 2016;3(5):415–24.

4. Cruz-Gonzalez P, He AWJ, Lam EPP, Ng IMC, Li MW, Hou R, et al. Artificial intelligence in mental health care: a systematic review of diagnosis, monitoring, and intervention applications. Psychol Med. 2025;55:e18.

5. Wanjau MN, Möller H, Haigh F, Milat A, Hayek R, Lucas P, et al. Physical Activity and Depression and Anxiety Disorders: A Systematic Review of Reviews and Assessment of Causality. AJPM Focus. 2023;2(2):100074.

6. Zhang Y, Stewart C, Ranjan Y, Conde P, Sankesara H, Rashid Z, et al. Large-scale digital phenotyping: Identifying depression and anxiety indicators in a general UK population with over 10,000 participants. J Affect Disord. 2025;375:412–22.

7. Gullett N, Zajkowska Z, Walsh A, Harper R, Mondelli V. Heart rate variability (HRV) as a way to understand associations between the autonomic nervous system (ANS) and affective states: A critical review of the literature. Int J Psychophysiol. 2023;192:35–42.

8. Sameh A, Nauha L, Seppänen M, Niemelä M, Oussalah M, Korpelainen R, et al. Digital phenotyping using wearable-determined physical behaviours and machine learning to detect depression and anxiety symptoms in a general population. Psychiatry Res. 2026;364:117242.

9. Correia ATL, Lipinska G, Rauch HGL, Forshaw PE, Roden LC, Rae DE. Associations between sleep-related heart rate variability and both sleep and symptoms of depression and anxiety: A systematic review. Sleep Med. 2023;101:106–17.

10. Hu MX, Milaneschi Y, Lamers F, Nolte IM, Snieder H, Dolan C V., et al. The association of depression and anxiety with cardiac autonomic activity: The role of confounding effects of antidepressants. Depress Anxiety. 2019;36:1163.

11. Chang H-A, Chang C-C, Chen C-L, Kuo TBJ, Lu R-B, Huang S-Y. Major depression is associated with cardiac autonomic dysregulation. Acta Neuropsychiatr. 2012;24:318–27.

12. Koch C, Wilhelm M, Salzmann S, Rief W, Euteneuer F. A meta-analysis of heart rate variability in major depression. Psychol Med. 2019;49(12):1948–57.

13. Assaad RH, Mohammadi M, Poudel O. Developing an intelligent IoT-enabled wearable multimodal biosensing device and cloud-based digital dashboard for real-time and comprehensive health, physiological, emotional, and cognitive monitoring using multi-sensor fusion technologies. Sensors Actuators A Phys. 2025;381:116074.

14. Sameh A, Rostami M, Oussalah M, Korpelainen R, Farrahi V. Digital phenotypes and digital biomarkers for health and diseases: a systematic review of machine learning approaches utilizing passive non-invasive signals collected via wearable devices and smartphones. Artif Intell Rev. 2025;58:1–43.

15. Roos LG, Slavich GM. Wearable technologies for health research: Opportunities, limitations, and practical and conceptual considerations. Brain Behav Immun. 2023;113:444–52.

16. Heckler WF, Feijó LP, de Carvalho JV, Barbosa JLV. Digital phenotyping for mental health based on data analytics: A systematic literature review. Artif Intell Med. 2025;163:103094.

17. Aboagye NY, Hinchliffe C, Del Din S, Ng W-F, Baker KF, Baker MR. Systematic review: digital biomarkers of fatigue in chronic diseases. npj Digit Med. 2025;8(1):602.

18. Mahato K, Saha T, Ding S, Sandhu SS, Chang AY, Wang J. Hybrid multimodal wearable sensors for comprehensive health monitoring. Nat Electron. 2024;7:735–50.

19. Sun Y, Chen J, Ji M, Li X. Wearable Technologies for Health Promotion and Disease Prevention in Older Adults: Systematic Scoping Review and Evidence Map. J Med Internet Res. 2025;27:e69077.

20. K, H., Rege, M., Paprika, Z.Z., Kumar, K.V.R., & Ganie SM. Smart Rings: Redefining Wearable Technology. Handb Artif Intell Wearables Appl Case Stud. 2024; p.217–35.

21. Herberger S, Aurnhammer C, Bauerfeind S, Bothe T, Penzel T, Fietze I. Performance of wearable finger ring trackers for diagnostic sleep measurement in the clinical context. Sci Rep. 2025;15(1):1–14.

22. Cao R, Azimi I, Sarhaddi F, Niela-Vilen H, Axelin A, Liljeberg P, et al. Accuracy Assessment of Oura Ring Nocturnal Heart Rate and Heart Rate Variability in Comparison With Electrocardiography in Time and Frequency Domains: Comprehensive Analysis. J Med Internet Res. 2022;24:e27487.

23. De Zambotti M, Cellini N, Goldstone A, Colrain IM, Baker FC. Wearable Sleep Technology in Clinical and Research Settings. Med Sci Sports Exerc. 2019;51(7):1538–57.

24. Kristiansson E, Fridolfsson J, Arvidsson D, Holmäng A, Börjesson M, Andersson-Hall U. Validation of Oura ring energy expenditure and steps in laboratory and free-living. BMC Med Res Methodol. 2023;23(1):1–11.

25. Fudolig MI, Bloomfield LSP, Price M, Bird YM, Hidalgo JE, Kim JN, et al. The Two Fundamental Shapes of Sleep Heart Rate Dynamics and Their Connection to Mental Health in College Students. Digit Biomarkers. 2024;8(1):120–31.

26. Norful AA, de Jacq K, Zhao J, Gao Y, Asadoorian K, Yang Y, et al. Exploring longitudinal physiologic stress measurement and sleep quality interventions to improve psychological well-being in nurses: a pilot study. Heal Psychol Behav Med. 2025;13(1) 2503376.

27. Torous J, Linardon J, Goldberg SB, Sun S, Bell I, Nicholas J, et al. The evolving field of digital mental health: current evidence and implementation issues for smartphone apps, generative artificial intelligence, and virtual reality. World Psychiatry. 2025;24(2):156.

28. University of Oulu. Northern Finland Birth Cohort 1986. 2020. http://urn.fi/urn:nbn:fi:att:f5c10eef-3d25-4bd0-beb8-f2d59df95b8e.

29. Miettunen J, Haapea M, Björnholm L, Huhtaniska S, Juola T, Kinnunen L, et al. Psychiatric research in the Northern Finland Birth Cohort 1986 – a systematic review. Int J Circumpolar Health. 2019;78 (1):1571382.

30. Järvelin MR, Elliott P, Kleinschmidt I, Martuzzi M, Grundy C, Hartikainen AL, et al. Ecological and individual predictors of birthweight in a northern Finland birth cohort 1986. Paediatr Perinat Epidemiol. 1997;11(3):298–312.

31. Kinnunen H, Rantanen A, Kenttä T, Koskimäki H. Feasible assessment of recovery and cardiovascular health: accuracy of nocturnal HR and HRV assessed via ring PPG in comparison to medical grade ECG. Physiol Meas. 2020;41(4):04NT01.

32. Birrer V, Elgendi M, Lambercy O, Menon C. Evaluating reliability in wearable devices for sleep staging. npj Digit Med 2024;7:1–14.

33. Alzueta E, de Zambotti M, Javitz H, Dulai T, Albinni B, Simon KC, et al. Tracking Sleep, Temperature, Heart Rate, and Daily Symptoms Across the Menstrual Cycle with the Oura Ring in Healthy Women. Int J Womens Health. 2022;14:491–503.

34. Ventevogel P, De Vries G, Scholte WF, Shinwari NR, Faiz H, Nassery R, et al. Properties of the Hopkins Symptom Checklist-25 (HSCL-25) and the Self-Reporting Questionnaire (SRQ-20) as screening instruments used in primary care in Afghanistan. Soc Psychiatry Psychiatr Epidemiol. 2007;42(4):328–35.

35. Cruz-Gonzalez M, Shrout PE, Alvarez K, Hostetter I, Alegría M. Measurement Invariance of Screening Measures of Anxiety, Depression, and Level of Functioning in a US Sample of Minority Older Adults Assessed in Four Languages. Front Psychiatry. 2021;12:579173.

36. Mattisson C, Bogren M, Horstmann V. Correspondence between clinical diagnoses of depressive and anxiety disorders and diagnostic screening via the Hopkins Symptom Check List-25 in the Lundby Study. Nord J Psychiatry. 2013;67(3):204–13.

37. Spitzer RL, Kroenke K, Williams JBW, Löwe B. A Brief Measure for Assessing Generalized Anxiety Disorder: The GAD-7. Arch Intern Med. 2006;166(10):1092–7.

38. Veijola J, Jokelainen J, Läksy K, Kantojärvi L, Kokkonen P, Järvelin M, et al. The Hopkins Symptom Checklist-25 in screening DSM-III-R axis-I disorders. Nord J Psychiatry. 2003;57(2):119–23.

39. Winokur A, Winokur DF, Rickels K, Cox DS. Symptoms of Emotional Distress in a Family Planning Service: Stability over a Four-Week Period. Br J Psychiatry. 1984;144:395–9.

40. Glaesmer H, Braehler E, Grande G, Hinz A, Petermann F, Romppel M. The German Version of the Hopkins Symptoms Checklist-25 (HSCL-25) — Factorial structure, psychometric properties, and population-based norms. Compr Psychiatry. 2014;55(2):396–403.

41. de Zambotti M, Rosas L, Colrain IM, Baker FC. The Sleep of the Ring: Comparison of the ŌURA Sleep Tracker Against Polysomnography. Behav Sleep Med. 2019;17(2):124–36.

42. Adaimi R, Thigpen N, Clausel A, Gotlieb N, Patel K, de Zambotti M. Temporal Trajectories in Sleep, Temperature Trends, Cardiorespiratory, and Activity Metrics Measured via Oura Ring During Pregnancy: Large-Scale Observational Analysis. JMIR Mhealth Uhealth. 2025;13:e80213.

43. Bloomfield LSP, Fudolig MI, Kim J, Llorin J, Lovato JL, McGinnis EW, et al. Predicting stress in first-year college students using sleep data from wearable devices. PLOS Digit Heal. 2024;3(4):e0000473.

44. Airlie J, Forster A, Birch KM. An investigation into the optimal wear time criteria necessary to reliably estimate physical activity and sedentary behaviour from ActiGraph wGT3X+ accelerometer data in older care home residents. BMC Geriatr. 2022;22(1):136.

45. Herrmann SD, Barreira T V, Kang M, Ainsworth BE. How Many Hours Are Enough? Accelerometer Wear Time May Provide Bias in Daily Activity Estimates. J Phys Act Heal. 2013(5);10:742–9.

46. Fini NA, Holland AE, Bernhardt J, Burge AT. How Many Hours of Device Wear Time Are Required to Accurately Measure Physical Activity Post Stroke? International Journal of Environmental Research and Public Health. 2022;19(3):1191.

47. To QG, Stanton R, Schoeppe S, Doering T, Vandelanotte C. Differences in physical activity between weekdays and weekend days among U.S. children and adults: Cross-sectional analysis of NHANES 2011–2014 data. Prev Med Reports. 2022;28:101892.

48. Price GD, Heinz M V., Collins AC, Jacobson NC. Detecting major depressive disorder presence using passively-collected wearable movement data in a nationally-representative sample. Psychiatry Res. 2024;332:115693.

49. Kim H-Y. Statistical notes for clinical researchers: post-hoc multiple comparisons. Restor Dent Endod. 2015;40(2):172–6.

50. Clark-Carter D. Analysis of Covariance (Ancova). Psychol Press. 2009. doi:10.4324/9780203870709-30.

51. Scott KM, Wells JE, Angermeyer M, Brugha TS, Bromet E, Demyttenaere K, et al. Gender and the relationship between marital status and first onset of mood, anxiety and substance use disorders. Psychol Med. 2010;40(9):1495–505.

52. Luppino FS, de Wit LM, Bouvy PF, Stijnen T, Cuijpers P, Penninx BWJH, et al. Overweight, Obesity, and Depression: A Systematic Review and Meta-analysis of Longitudinal Studies. Arch Gen Psychiatry. 2010;67(3):220–9.

53. Fluharty M, Taylor AE, Grabski M, Munafò MR. The Association of Cigarette Smoking With Depression and Anxiety: A Systematic Review. Nicotine Tob Res. 2017;19(1):3–13.

54. Pan A, Keum N, Okereke OI, Sun Q, Kivimaki M, Rubin RR, et al. Bidirectional Association Between Depression and Metabolic Syndrome: A systematic review and meta-analysis of epidemiological studies. Diabetes Care. 2012;35(5):1171–80.

55. O’brien RM. A Caution Regarding Rules of Thumb for Variance Inflation Factors. Qual Quant. 2007;41:673–90.

56. statsmodels · PyPI. https://pypi.org/project/statsmodels/. Accessed 28 Oct 2025.

57. Meeus M, Goubert D, De Backer F, Struyf F, Hermans L, Coppieters I, et al. Heart rate variability in patients with fibromyalgia and patients with chronic fatigue syndrome: A systematic review. Semin Arthritis Rheum. 2013;43(2):279–87.

58. Zabara-Antal A, Crisan-Dabija R, Arcana R-I, Melinte OE, Pintilie A-L, Grosu-Creanga IA, et al. Heart Rate Variability (HRV) in Patients with Sleep Apnea and COPD: A Comprehensive Analysis. Journal of Clinical Medicine. 2025;14(13):4630.

59. Rama AN, Charles Cho S, Kushida CA. Chapter 2 NREM–REM sleep. In: Guilleminault CBT-H of CN, editor. Handbook of Clinical Neurophysiology. Vol. 6. Elsevier; 2005. p. 21–9.

60. Vasiliadis H-M, Chudzinski V, Gontijo-Guerra S, Préville M. Screening instruments for a population of older adults: The 10-item Kessler Psychological Distress Scale (K10) and the 7-item Generalized Anxiety Disorder Scale (GAD-7). Psychiatry Res. 2015;228(1):89–94.

61. Kwon N, Lee D, Hwang S, Yang S, Youn I, Moon HJ, et al. Enhancing the accuracy of mental health assessments through the integration of self-report and objective measures: A convergence study utilizing biosignals and 14-day wearable data. Acta Psychol (Amst). 2025;259:105432.

62. Rykov Y, Thach TQ, Bojic I, Christopoulos G, Car J. Digital biomarkers for depression screening with wearable devices: Cross-sectional study with machine learning modeling. JMIR mHealth uHealth. 2021;9(10):e24872.

63. Zhang Y, Folarin AA, Sun S, Cummins N, Ranjan Y, Rashid Z, et al. Longitudinal Assessment of Seasonal Impacts and Depression Associations on Circadian Rhythm Using Multimodal Wearable Sensing: Retrospective Analysis. J Med Internet Res. 2024;26:e55302.

64. Agorastos A, Mansueto AC, Hager T, Pappi E, Gardikioti A, Stiedl O. Heart Rate Variability as a Translational Dynamic Biomarker of Altered Autonomic Function in Health and Psychiatric Disease. Biomedicines. 2023;11(6):1591.

65. Ferreira JJ, Fernandes CI, Rammal HG, Veiga PM. Wearable technology and consumer interaction: A systematic review and research agenda. Comput Human Behav. 2021;118:106710.

66. Guo X, Su T, Xiao H, Xiao R, Xiao Z. Using 24-h Heart Rate Variability to Investigate the Sleep Quality and Depression Symptoms of Medical Students. Front Psychiatry. 2022;12:781673.

67. Lau Y, Chemas N, Ajeet Gokani H, Morrell R, Phannarus H, Cooper C, et al. Association Between Digital Biomarkers of Health and Anxiety: Systematic Review and Meta-Analysis. J Med Internet Res. 2026;28:e73812.

68. Tomasi J, Zai CC, Zai G, Herbert D, Richter MA, Mohiuddin AG, et al. Investigating the association of anxiety disorders with heart rate variability measured using a wearable device. J Affect Disord. 2024;351:569–78.

69. Kerkering EM, Greenlund IM, Bigalke JA, Migliaccio GCL, Smoot CA, Carter JR. Reliability of heart rate variability during stable and disrupted polysomnographic sleep. Am J Physiol Circ Physiol. 2022;323(1):H16–23.

70. de Zambotti M, Goldstein C, Cook J, Menghini L, Altini M, Cheng P, et al. State of the science and recommendations for using wearable technology in sleep and circadian research. Sleep. 2024;47(4):1–31.

71. Shaffer F, Ginsberg JP. An Overview of Heart Rate Variability Metrics and Norms. Front Public Heal. 2017;5:290215.

72. Chalmers JA, Quintana DS, Abbott MJA, Kemp AH. Anxiety disorders are associated with reduced heart rate variability: A meta-analysis. Front Psychiatry. 2014;5 JUL:100467.

73. Jo YT, Lee SW, Park S, Lee J. Association between heart rate variability metrics from a smartwatch and self-reported depression and anxiety symptoms: a four-week longitudinal study. Front Psychiatry. 2024;15:1371946.

74. Siddi S, Bailon R, Giné-Vázquez I, Matcham F, Lamers F, Kontaxis S, et al. The usability of daytime and night-time heart rate dynamics as digital biomarkers of depression severity. Psychol Med. 2023;53(8):3249–60.

75. Barak O, Bauer AD, Vaisbuch E, Piekos SN, Sadovsky Y. Wearable-based phenotyping of activity patterns during pregnancy. Pregnancy. 2026;2:e70326.

76. Daniore P, Nittas V, Haag C, Bernard J, Gonzenbach R, von Wyl V. From wearable sensor data to digital biomarker development: ten lessons learned and a framework proposal. npj Digit Med. 2024;7(1):161.

77. Strain T, Wijndaele K, Dempsey PC, Sharp SJ, Pearce M, Jeon J, et al. Wearable-device-measured physical activity and future health risk. Nat Med. 2020;26 (9):1385–91.

78. Moshe I, Terhorst Y, Opoku Asare K, Sander LB, Ferreira D, Baumeister H, et al. Predicting Symptoms of Depression and Anxiety Using Smartphone and Wearable Data. Front Psychiatry. 2021;12:625247.

79. Fiore M, Bianconi A, Sicari G, Conni A, Lenzi J, Tomaiuolo G, et al. The Use of Smart Rings in Health Monitoring—A Meta-Analysis. Appl Sci. 2024;14(23):10778.

80. Sivertsen B, Skogen JC, Reneflot A, Knapstad M, Smith ORF, Aarø LE, et al. Assessing Diagnostic Precision: Adaptations of the Hopkins Symptom Checklist (HSCL-5/10/25) Among Tertiary-Level Students in Norway. Clin Psychol Eur. 2024;6:1–18.

81. Mehrabadi MA, Azimi I, Sarhaddi F, Axelin A, Niela-Vilén H, Myllyntausta S, et al. Sleep tracking of a commercially available smart ring and smartwatch against medical-grade actigraphy in everyday settings: Instrument validation study. JMIR mHealth uHealth. 2020;8(10):e20465.

82. Ellender CM, Zahir SF, Meaklim H, Joyce R, Cunnington D, Swieca J. Prospective cohort study to evaluate the accuracy of sleep measurement by consumer-grade smart devices compared with polysomnography in a sleep disorders population. BMJ Open. 2021;11(11):e044015.

83. Khan S, Ibrahim AF, Vasudevan SS, Quatela OE, Nanu DP, Carr MM. The Oura Ring Versus Medical-Grade Sleep Studies: A Systematic Review and Meta-Analysis. OTO Open. 2025;9 (4):e70181.

84. Wang Z, Luo Y, Zhang Y, Chen L, Zou Y, Xiao J, et al. Heart rate variability in generalized anxiety disorder, major depressive disorder and panic disorder: A network meta-analysis and systematic review. J Affect Disord. 2023;330:259–66.

